# Modeling the COVID-19 outbreak in Ecuador: Is it the right time to lift social distancing containment measures?

**DOI:** 10.1101/2020.05.21.20109520

**Authors:** Miguel Reina Ortiz, Vinita Sharma

## Abstract

**Objective:** Model the effect of partial and full reversal of containment measures on COVID-19 morbidity and mortality in Ecuador.

**Methods:** Susceptible, Infected, Recovered (SIR) models were used to simulate the transmission dynamics of COVID-19 before and after the implementation (and reversal) of containment measures. A Healthcare Compartmental Epidemic Model (HeCEM), which accounts for Hospital and Intensive Care Unit admission rates, was developed to also simulate the effect of reversing social distancing containment measures. Reported COVID-19 cases between February 29^th^ and April 23^rd^, 2020 were obtained from the Servicio Nacional de Gestión de Riesgos y Emergencia, Ecuador’s national emergency management office. An ARIMA model was used to forecast reported number of cases based on the reported number of cases. SIR, HeCEM, and ARIMA model prediction errors were estimated.

**Results:** SIR and HeCEM models predict that, at the moment, hospital and ICU bed needs for COVID-19 patients exceed the capacity in Ecuador. Partial or full reversal of containment measures before reaching the point where hospital and ICU beds are enough to meet the expected demand will result in secondary waves that delay reaching this equilibrium, resulting in thousands of excess deaths. Forecasts predict over 50,000 reported COVID-19 cases by July 25^th^, 2020.

**Conclusion:** Partial reversal of containment measures should occur only after enough hospital and beds are available to meet the demand.

## INTRODUCTION

According to the World Health Organization (WHO), almost 4 million COVID-19 cases and more than 250,000 COVID-19-related deaths had been reported around the world as of May 10, 2020. (1) With over 28,000 cases, Ecuador ranks sixth in terms of absolute number of cases in the Americas. (1) However, when accounting for population size, Ecuador ranks third (first among Latin American countries). Thus, Ecuador has become a regional COVID-19 pandemic epicenter for the Latin American region.

Ecuador reported its first case of COVID-19 on February 29^th^, 2020, (2). The country declared a State of Sanitary Emergency on March 11^th^, 2020, which entailed suspension of school activities and a ban on gatherings of ≥ 250 people. (3) Since March 17^th^, Ecuador has increased social distancing measures by progressively restricting in-country mobility. (4, 5) Restrictions implemented since March 18^th^ include: 1) suspension of intercity transport; (5) 2) reduction of inner-city car circulation;(5) and, 3) imposing a curfew, (5) among others.

In recent days, local governments have been given the authority to decide when and how to move from more restrictive “red” to less restrictive “green” phases in a 3-phased system resembling traffic lights. (6) The objective of this article is to provide evidence to inform decisions on reversing containment measures by conducting an epidemiological description of Ecuador’s COVID-19 outbreak, by modeling the effect of partial reversal of containment measures under different scenarios, and by forecasting predicted number of new reported COVID-19 cases in the country.

## METHODS

### Study Population

Ecuador is a country located in the northwest of South America. It borders Colombia to the north, Peru to the east and south, and the Pacific Ocean lies to its west. Ecuador also shares a maritime border with Costa Rica. The population of Ecuador is estimated to be 17,084,357 in 2020. (7) The country is administratively and politically subdivided into provinces, cantons, and parishes. There are 24 provinces and 221 cantons (8) in Ecuador.

### Study design

Based on secondary data analysis, an ecological study was conducted to describe the epidemiological characteristics of Ecuador’s COVID-19 outbreak, including model-generated predictions and forecasts of future incidence and deaths.

### Data collection

Cumulative number of COVID-19 cases and cumulative number of COVID-19 deaths (per day) were obtained from the COVID-19 reports published by the National Emergency Operations Committee (*COE Nacional*, in Spanish), Servicio Nacional de Gestion de Riesgos (National Service of Risk Management), Government of Ecuador. (9) Based on this data, we constructed a database of cumulative COVID-19 cases and deaths from Day 0 of the outbreak (February 29^th^, 2020) (2) until April 23^rd^, 2020. Data collection stopped at April 23^rd^, 2020 because more than 10,000 cases (almost a 100% increase from the previous day) were added on April 24^th^, 2020 due to the inclusion of antibody-based test results, which had not been reported until then (i.e. previous days were based on RT-PCR results).

### Data Analysis

#### Epidemic Models – SIR

Compartmental deterministic models were simulated using the deSolve package(10), R software (11), to compare the potential impact of reversing containment measures on COVID-19 morbidity and mortality. Previous research has suggested that Susceptible, Infected, Recovered (SIR) models are more appropriate for modeling COVID-19 than Susceptible, Exposed, Infected, Recovered (SEIR) models. (12)

We simulated SIR Models where the total population of Ecuador (as reported by the World Bank) (7) was divided into Susceptible (S), Infectious (I), and Recovered(R) compartments. Ecuador reported its first case of COVID-19 in February 29^th^, 2020, (2) and this was selected as the starting date of the outbreak for our models. We model the outbreak without any containment measures (Model 1), the effect of the enhanced social distancing measures enacted on Mary 18^th^ (Model 2). We also model the effect of full or partial reversal of current social distancing measures at weekly intervals (on Sundays), starting on May 17^th^, 2020. Model parameters were derived from the published literature. (13)

For each scenario, we found the time at which the demand of hospital and ICU beds by COVID-19 patients no longer exceeds the capacity in Ecuador (i.e. after the outbreak peak). We call this point *Post-peak Hospital* and *Post-Peak Intensive Care Unit* (*ICU*) *Capacitance*. The number of hospital and Intensive Care Unit (ICU) beds available were obtained from the National Institute of Statistics and Census. (14) Hospital and ICU beds needed at any given point of time were calculated from SIR model outputs using a 20% and a 2% ratio, respectively. (15) The number of deaths at any given point of time was also calculated from SIR outputs by using a 2% case fatality ratio. (15)

#### Epidemic models (Healthcare)

Since SIR models do not account for the time that an individual is likely to remain in a hospital or ICU unit until either full recovery or death, we developed the compartmental model shown in Figure 1. In this model, susceptible people become infectious at a rate β (i.e. transmission rate). Infectious people can move into four different compartments: 1) recovery, at a rate *γ*; 2) hospitalization, at a rate θ; 3) ICU unit, at a rate ι; or, 4) deceased, at a rate δ. Hospitalized patients either recover at a rate ε or pass away at a rate χ whereas patients admitted to the ICU recovers at a rate ω or pass away at a rate α. Of note, the model allows to account for hospital-acquired infections, which occur at a rate ζ when hospitalized patients interact with healthcare workers. Once infected, a healthcare worker moves to the infectious compartment. The model also accounts for infections that healthcare workers may acquire from interaction with infectious, but not hospitalized, individuals (see Figure 1). Finally, the model estimates the number of reported cases (i.e. “observed” cases) based on a reporting rate μ (not shown in Figure 1).

**Figure 1.**
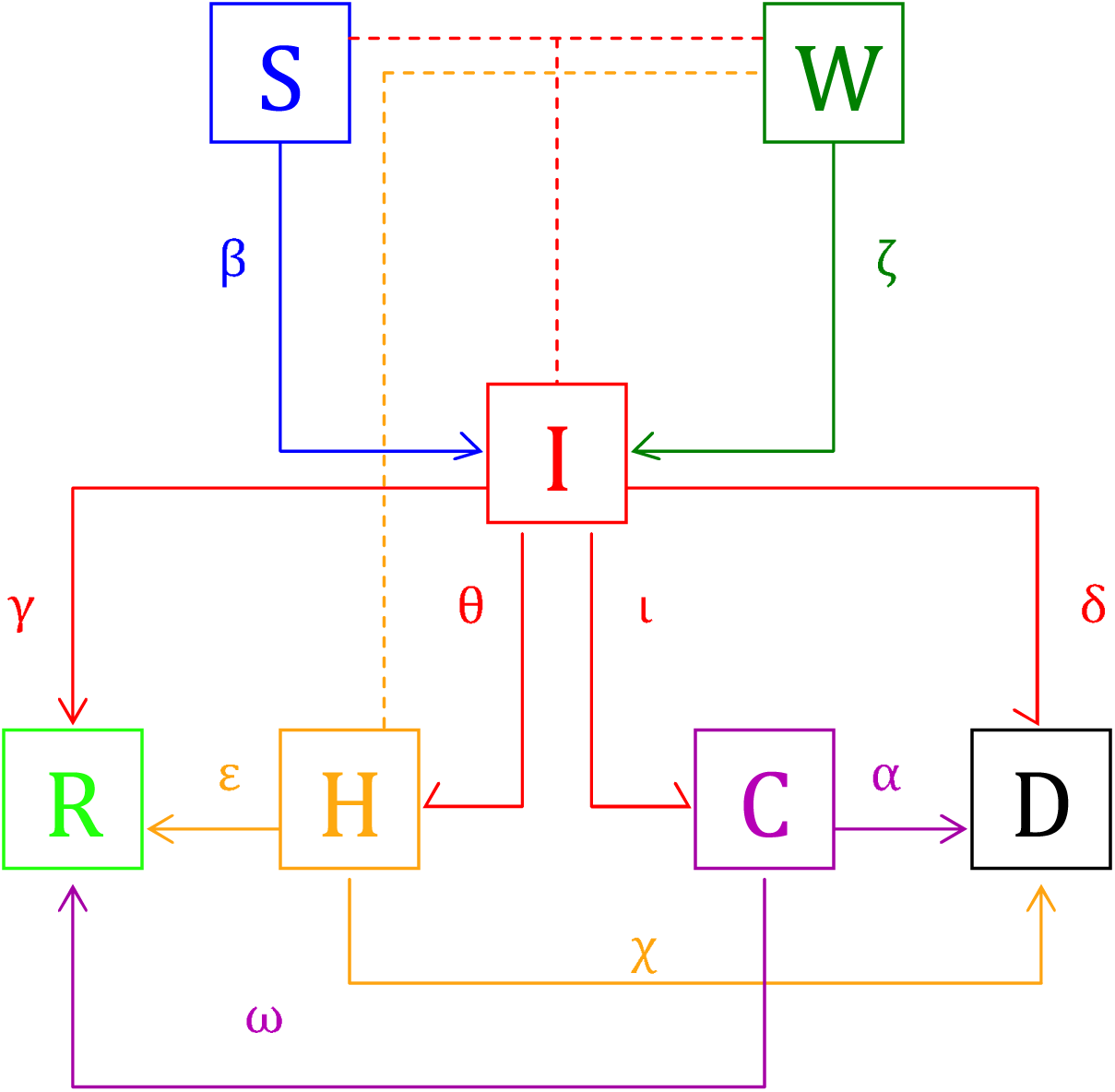
Healthcare Compartmental Epidemic Model. S = Susceptibles, W = Heatlhcare Workers, I = Infectious, R = Recovered, H = Hospitalized, C = admitted to ICU units, D = deceased. Greek letters represent the rate at which individuals move from one compartment to another. Arrows represent the direction of the inter-compartmental movement. Dashed lines represent interactions between people in different compartments that may result in infections.

The Healthcare Compartmental Epidemic Model consists of the following structure:

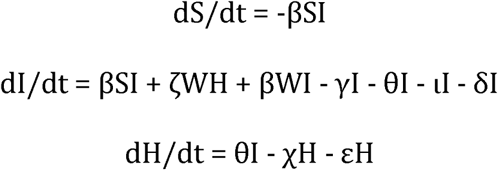

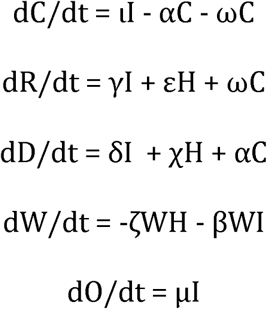

Parameters were derived from the published literature (Table 1).

**Table 1.** Healthcare Compartmental Epidemic Model Parameters.

| Parameter | Range | Period |
| --- | --- | --- |
| Infectious Period (before containment measures) | -- | 3.48 days(13) |
| Infectious Period (after containment measures) | -- | 3.13 days(13) |
| Hospitalization ratio | 6.8%* (16)to<br>31.4%(15)** | 7 days(17, 18) |
| ICU admission ratio<sup>+</sup> | 32% | 3.5 days(18) |
| Case fatality ratio (CFR) | 1.4%(16) to<br>3.4%(15)<sup>\$</sup> | 12.8 days(16) |
| CFR in ICU-admitted patients | 38%(18) | 7.3 days(16) |
| CFR in hospitalized patients | 4%(18) | 10.8 days(16) |
| Recovery ratio in ICU-admitted patients | 62%(18) | 17.7 days(16) |
| Recovery ratio in hospitalized patients | 96%(18) | 17.7 days(16) |

\* Weighted average, \*\* Other estimates: 12.1%(17), 20.7%,(15) 14%(19).
<sup>+</sup> among hospitalized patients.
<sup>\$</sup> Other estimates: 1.8%(15)
CFR = Case Fatality Ratio; ICU = Intensive Care Unit

The total number of healthcare workers was obtained from the National Institute of Statistics and Census. (20) We included the number of physicians, nurses and nurse assistants and excluded dentists, psychologists and midwives, as the latter were deemed less likely to enter in direct contact with a COVID-19 patient during the early pandemic spread.

#### Forecasting

ARIMA models were used to forecast cumulative COVID-19 reported incidence (i.e. number of reported cases by the system) beyond May 4^th^, 2020. In order to avoid bias introduced by the change in method of reporting, the forecast will be based on RT-PCR results, and only RT-PCR-confirmed cases will be forecasted. Optimal order of differencing was determined by examining autocorrelation function (ACF) and partial autocorrelation function (PACF) plots (Figure S1) and by comparing root mean square error (RMSE) terms of the fitted models (0,1, and 2 orders of differencing were tested, at this point RMSE increased without improving ACF and PACF plots). An inspection of the differentiated ACF and PACF plots revealed a slightly significant positive autocorrelation at lag-1 (Figure SI). Consequently, a (first-order) differenced first-order autoregressive model was fitted, controlling for outliers (AR parameter = 0.725, p < 0.001; Table S5) to forecast cumulative reported incidence beyond May 4^th^ (the date when the 3-phase traffic light system was implemented). Forecast was stopped on July 25^th^, 2020. ARIMA models were fit using SPSS 25. (21)

#### Simulated and Predicted errors

Simulated (SIR and HeCEM models) and predicted (ARIMA model) errors were used to compare observed against simulated/predicted cumulative incidence. In order to account for heteroscedasticity in the simulated/predicted errors, standardized errors were computed. Afterwards, one-sample T (or Wilcoxon signed-rank tests, as appropriate) analyses were conducted to assess whether error means (or medians) were different than zero.

## RESULTS

### Epidemiological characteristics of COVID-19 in Ecuador

In Ecuador, the number of reported cases has increased at a mean rate of 25% (SD = 68.03; Figure 2). The weekly mean increase rate shows a steady but non-monotonic decrease with the last two weeks showing a rate lower than 5% compared to over 80% on the first week (Table S1). As of April 23^rd^, the cumulative COVID-19 incidence curve still shows an upward trend with a total of 11,183 cases, a 3.07% increase from the previous day.

**Figure 2.**
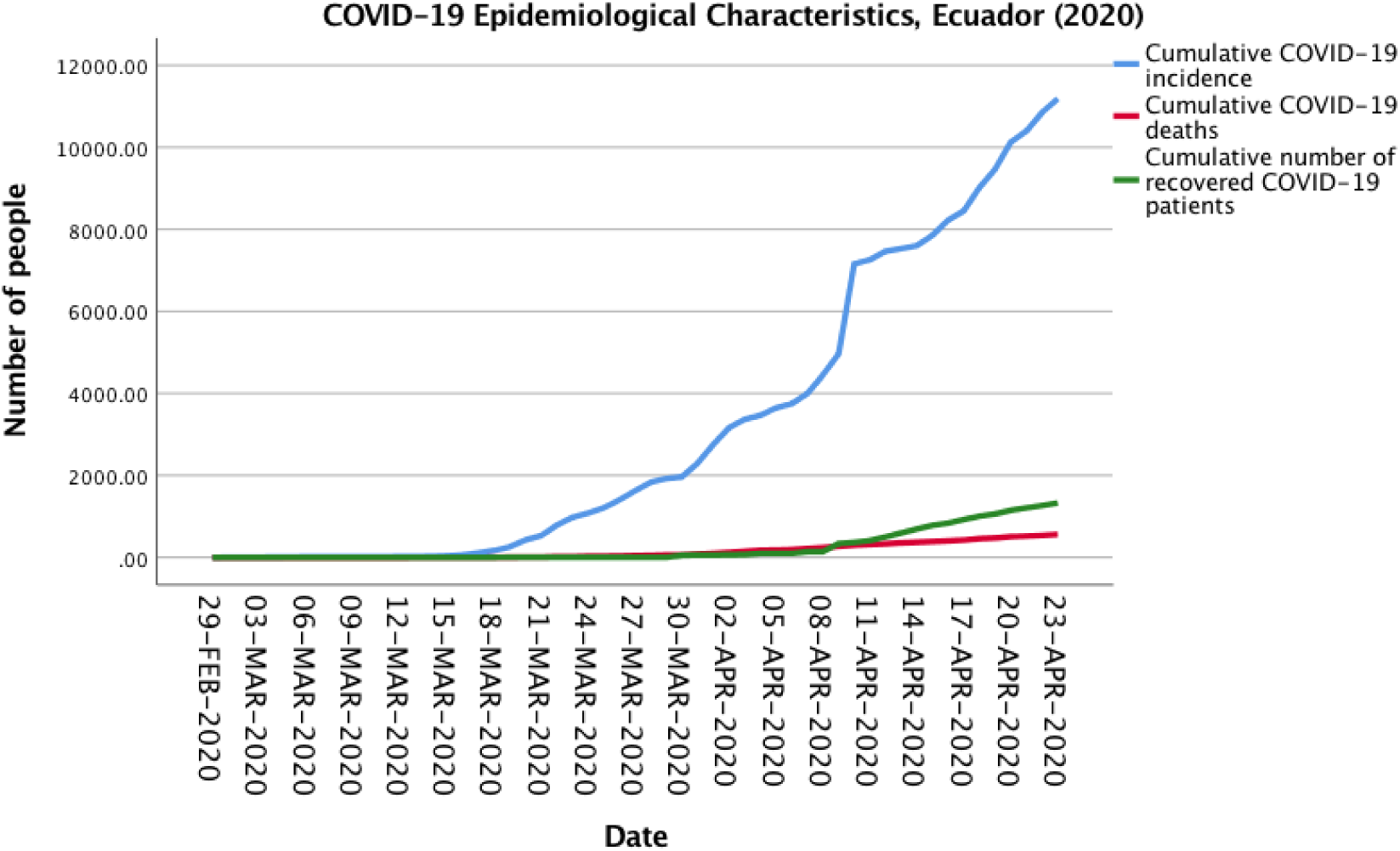
Epidemiological characteristics of the COVID-19 outbreak in Ecuador.

The country reported its first COVID-19 fatality on March 13^th^. (22) Since then, the cumulative number of deaths has increased at a mean rate of 18.61%. By April 23^rd^, there were 560 cumulative COVID-19 deaths reported. Although the mean number of reported deaths has remained relatively constant since Week 6, the rate of increase has progressively decreased from 22.62% on Week 4 to 4.29% on Week 9.

### Simulating the COVID-19 outbreak in Ecuador

Model 1 shows COVID-19 outbreak dynamics without accounting for the effect of containment measures (Figure 3A). In that context, the peak of the outbreak would have occurred on April 11^th^, 2020. Post-peak Hospital Capacitance would have been reached on May 2^nd^ whereas post-peak ICU Capacitance would have been reached by May 10^th^.

**Figure 3A.**
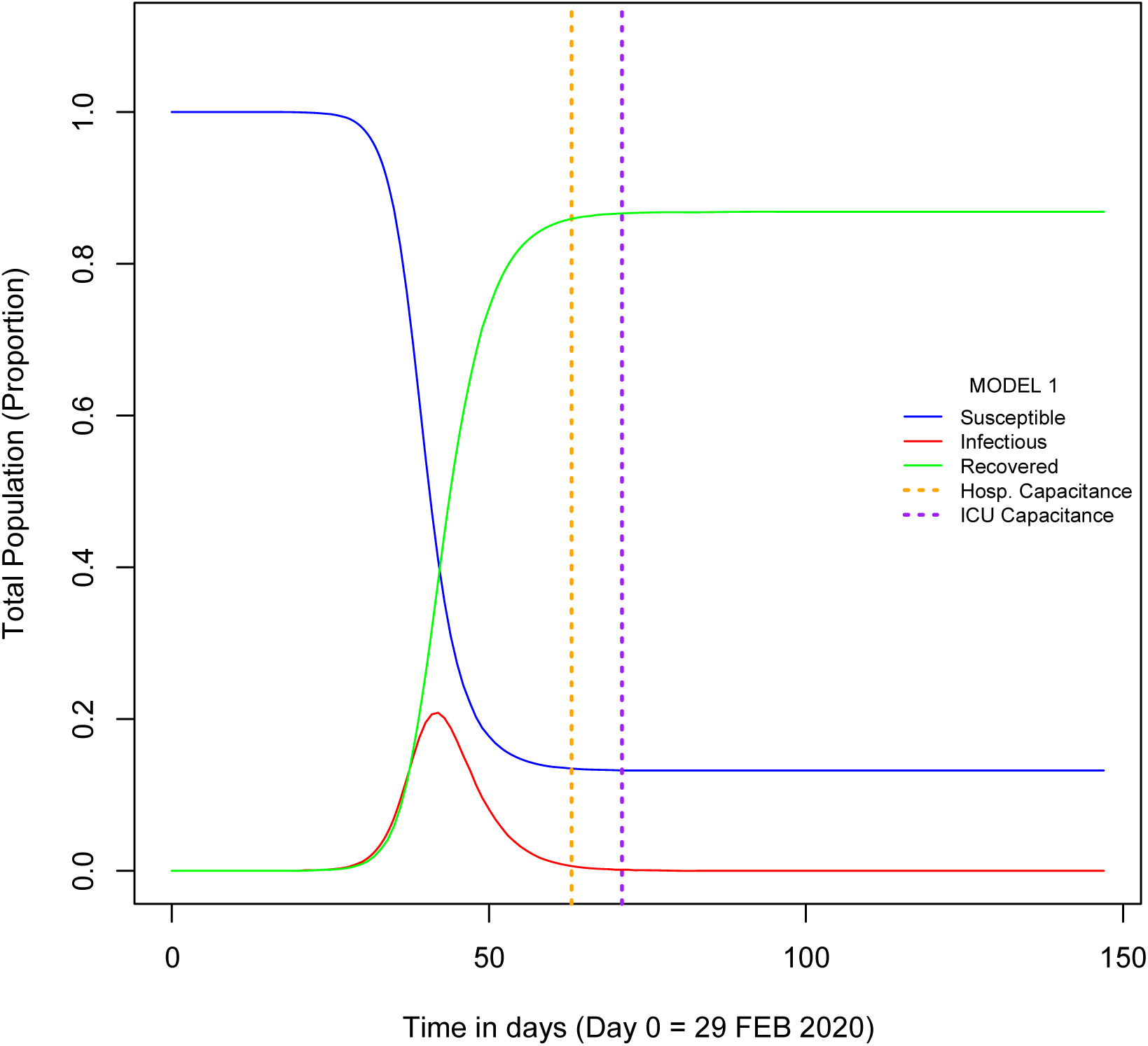
MODEL 1: Ecuador COVID-19 initial SIR Model.

Model 2 accounts for the restricting mobility social distancing measures imposed in Ecuador on March 18^th^, 2020 (Figure 3B). The time of the policy change (i.e. introduction of containment measures) effectively divides the model into Period A (before the implementation of such measures) and Period B (after). After accounting for the effect of containment measures, the peak of the COVID-19 outbreak is delayed until May 24^th^, 2020. Post-Peak Hospital Capacitance (HC) would be reached on June 2^nd^ whereas Post-peak ICU capacitance (ICU-C) would be reached by July 14^th^. As shown in Table S2, the introduction of containment measures is simulated to have reduced the number of new infections during the outbreak peak by 90% and to have averted almost 138,000 deaths.

### Impact of reversing containment measures on COVID-19 incidence

In order to analyze the temporal dynamics of the COVID-19 outbreak in Ecuador and to assess the impact that phasing out containment measures has on simulated COVID-19 morbidity and mortality we developed *full reversal* and *partial reversal* scenarios. Four full reversal (Models 3 to 6, Figures S2 to S5) and four partial reversal scenarios (Models 7 to 10, Figures S6 to S8 and Figure 3C) are presented. For each scenario, we assume that there will be a complete (or partial) reversal of containment measures at different time points, which are designated as “Policy change 2”. Each scenario is divided into three periods: 1) Period A, between start of outbreak and “Policy change 1”; 2) Period B, between “Policy change 1” and “Policy change 2”; and, 3) Period C, after “Policy change 2”.

**Figure 3B.**
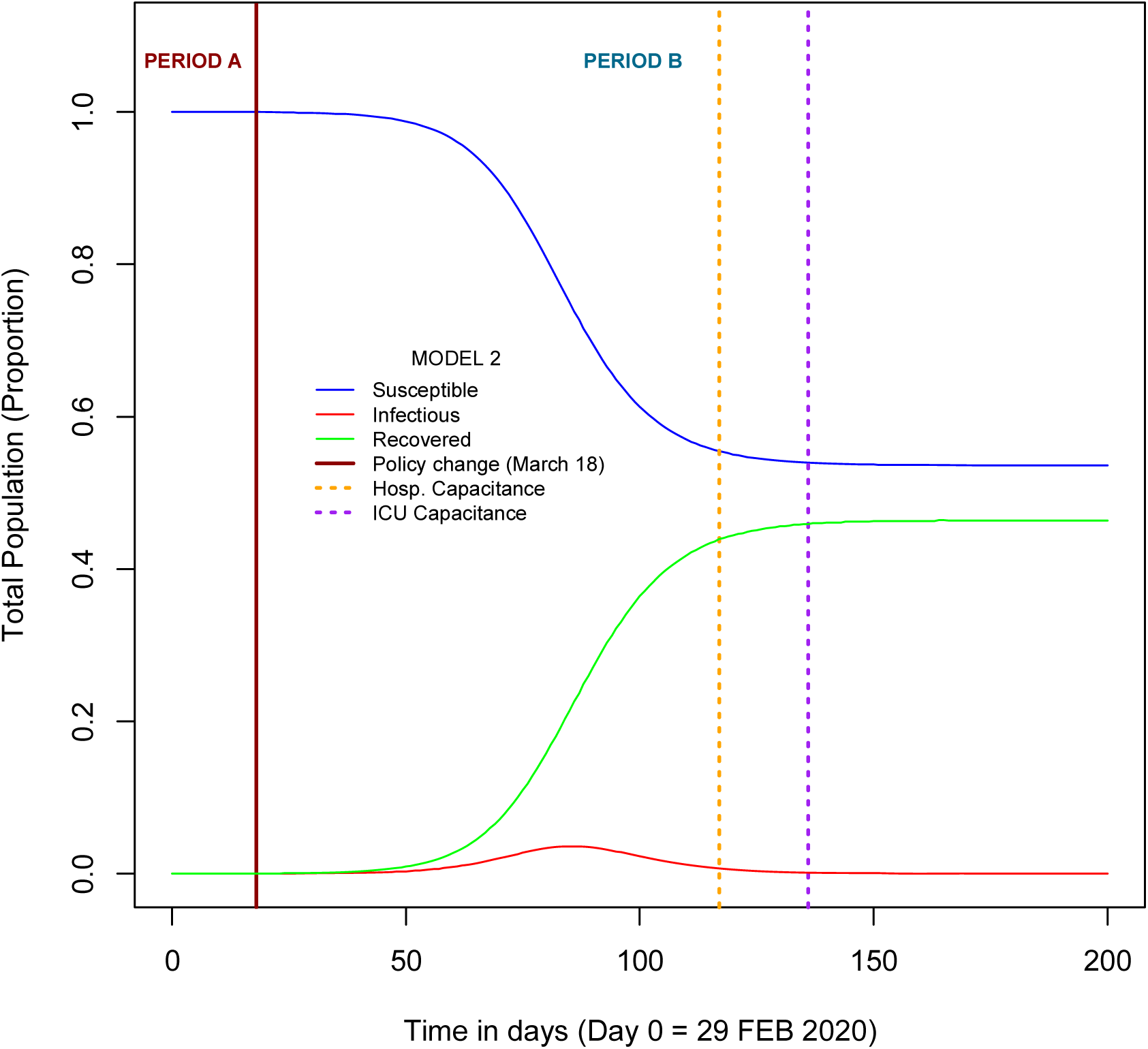
MODEL 2: SIR-simulated COVID-19 outbreak in Ecuador, accounting for the start of containment measures.

**Figure 3C.**
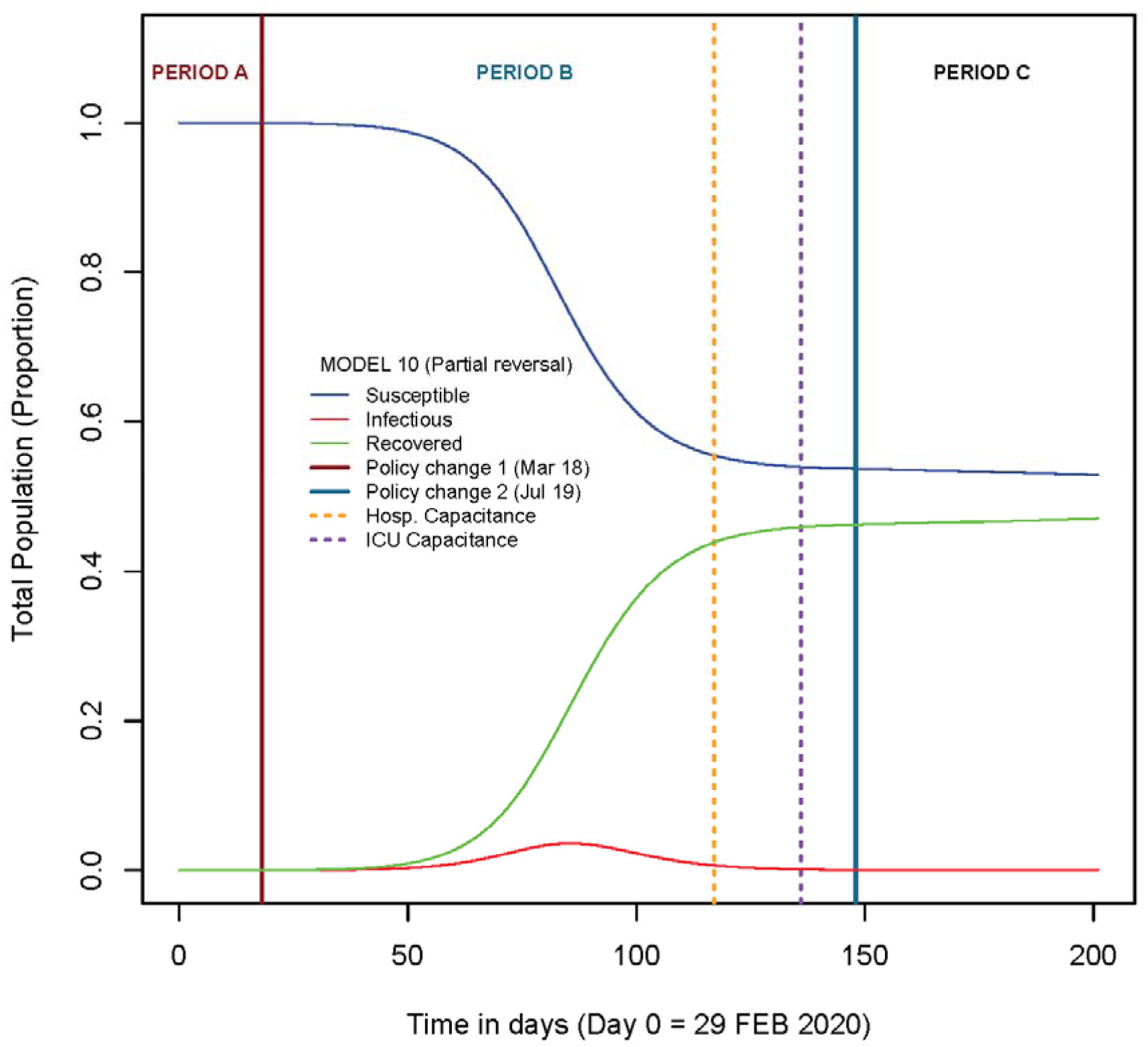
MODEL 10: SIR-simulated COVID-19 outbreak in Ecuador if containment measures are partially reversed on July 19^th^, 2020. Solid lines represent period boundaries. Dashed lines represent time at which capacitance is reached.

#### Full reversal scenarios

All full reversal scenarios would result in excess deaths ranging from 5,043 to 127,745 (Table S4). Only a full reversal on May 17^th^ would reach HC and ICU-C earlier than Model 2 (by 7 and 17 days, respectively. However, fully reversing social distancing containment measures by May 17^th^ would result in an excess of 127,745 deaths. More detailed descriptions for each individual scenario are provided in Supplemental Materials.

#### Partial reversal scenarios

Excess COVID-19 deaths range between 2,707 and 91,151. A partial reversal on May 17^th^ would result in HC and ICU-C being reached with 2 and 10 days of anticipation but with a cost of over 90,000 deaths, compared to keeping containment measures. The best scenario is provided by a partial reversal on July 9^th^, which would result in a 0- and 67-day delay for HC and ICU-C, compared with no change in policy (Model 10, Figure 3C). Excess mortality of a partial reversal on July 9^th^ would be 2,707 (Table S4). More detailed descriptions for each individual scenario are provided in Supplemental Materials.

### Simulating local COVID-19 outbreaks using HeCEM

We used HeCEM to find the predicted dates of HC and ICU-C for Guayaquil (Figure 4C, Model 13). ICU-C would be reach by June 30^th^ whereas HC would be reached on July 18^th^ (Figure 4C, Model 13). By May 10^th^, the model predicts over 1.1 million persons having been infected with COVID-19 in the city and about 15,000 deaths. In addition, it is predicted that by May 10^th^ over 6,500 healthcare workers have been infected with COVID-19 in Guayaquil.

**Figure 4A.**
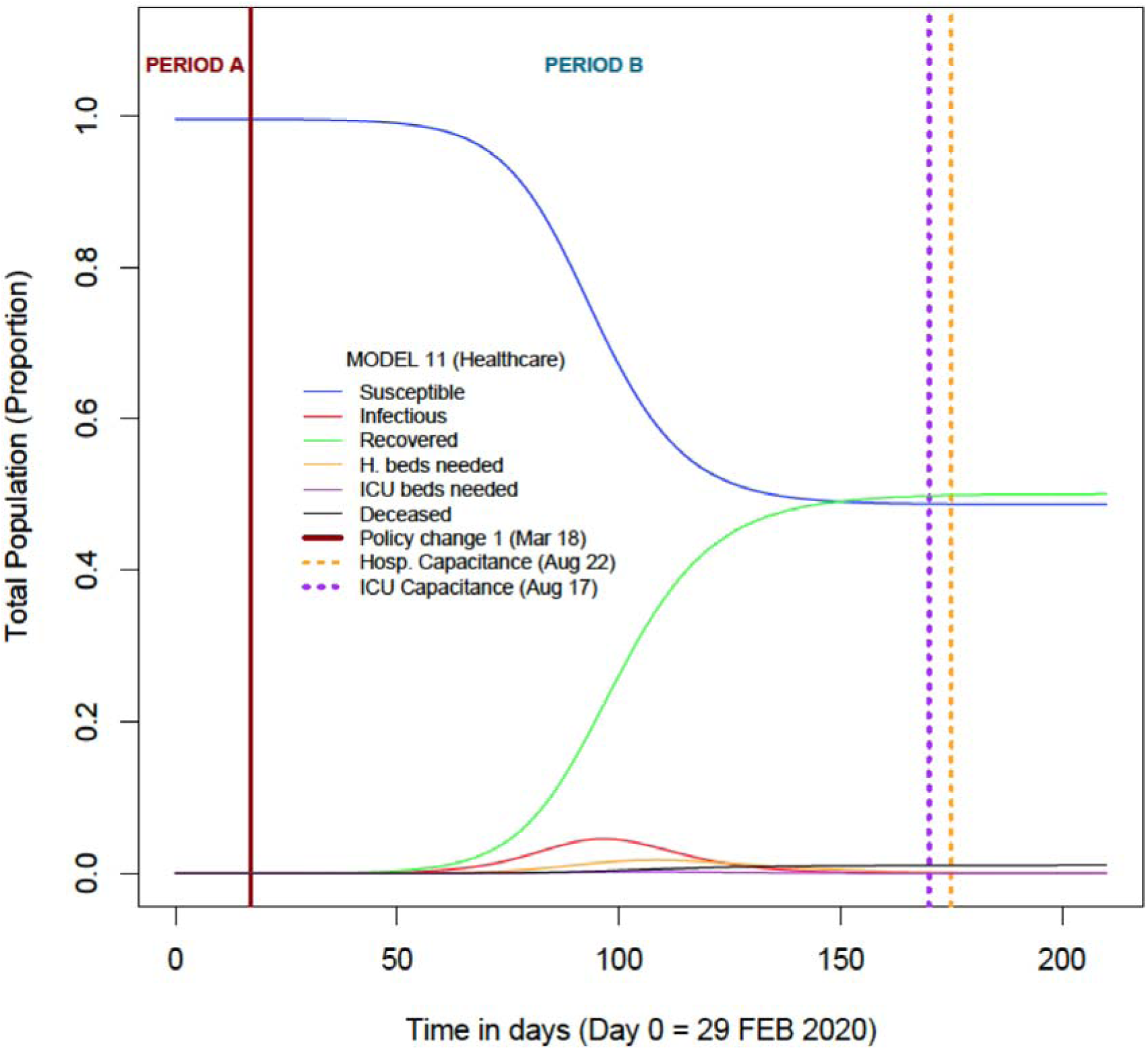
MODEL 11: HeCEM-simulated COVID-19 outbreak in Ecuador. Solid line represents the time when containment measures were enacted. Dashed lines represent time at which capacitance is reached.

**Figure 4B.**
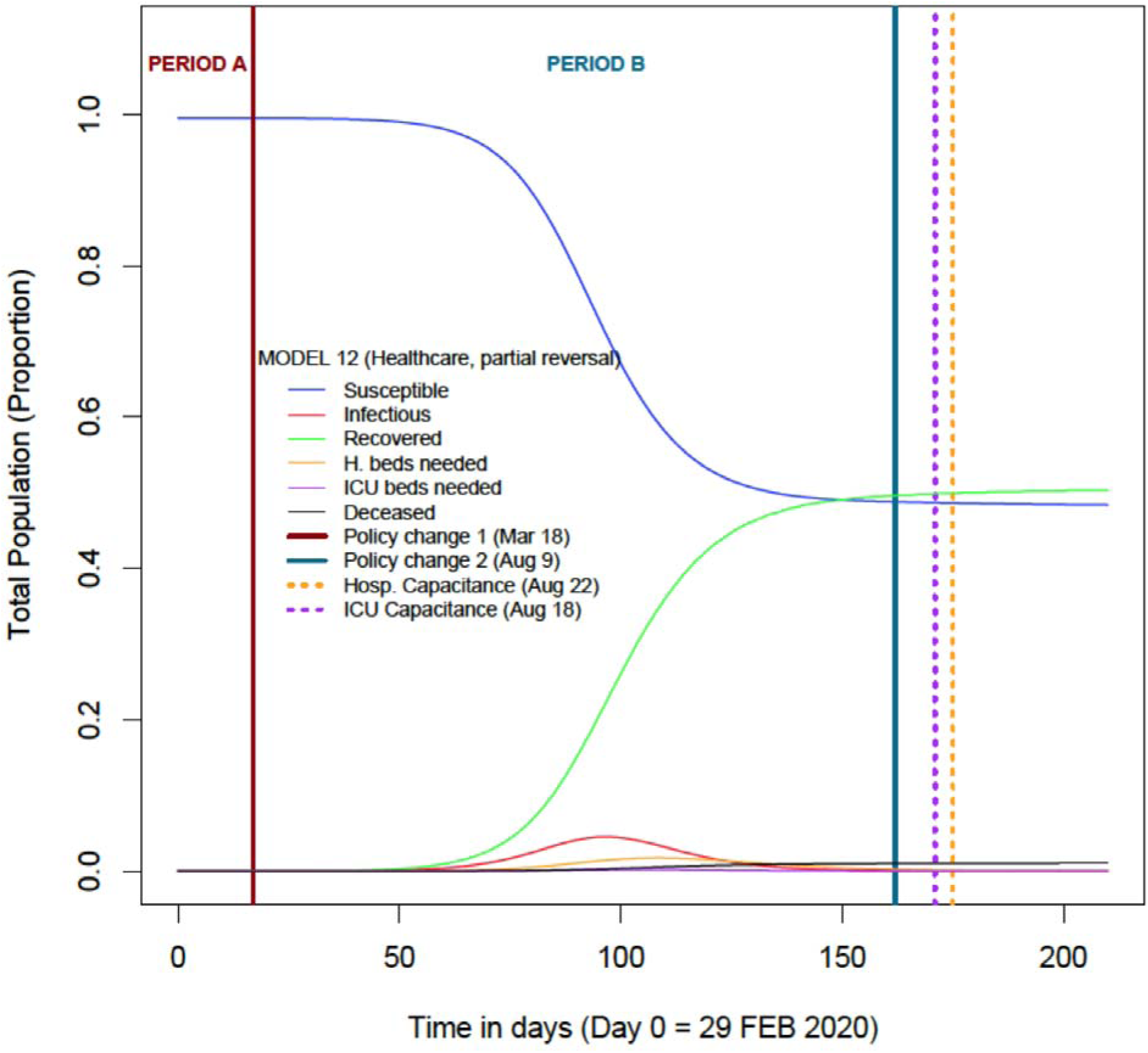
MODEL 12: HeCEM-simulated COVID-19 outbreak in Ecuador assuming a partial reversal of containment measures on August 9^th^. Solid lines represent time when COVID-19 policies change. Dashed lines represent time at which capacitance is reached.

**Figure 4C.**
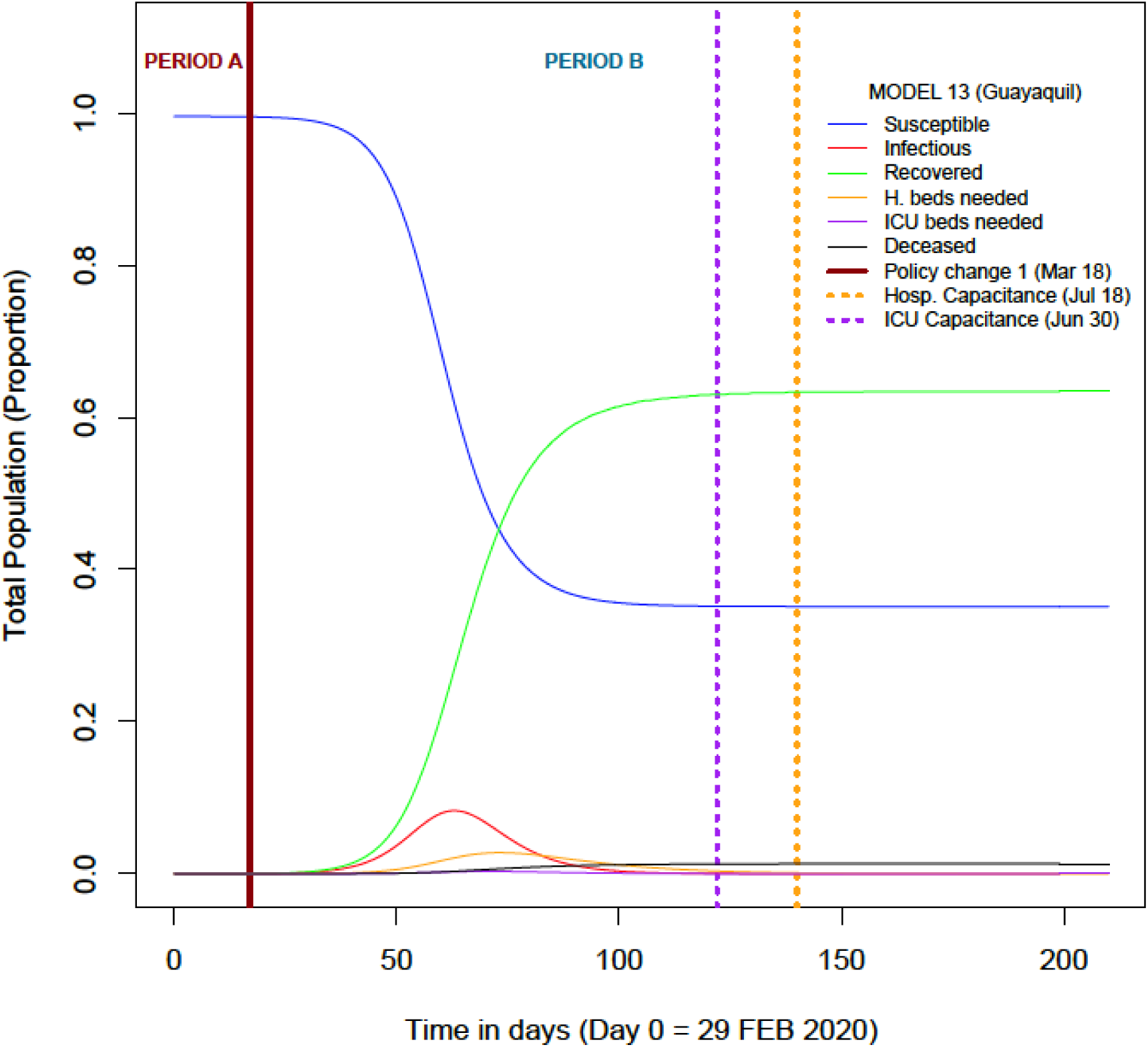
MODEL 13: HeCEM-simulated COVID-19 outbreak in Guayaquil. Solid lines represent time when COVID-19 policies change. Dashed lines represent time at which capacitance is reached.

**Figure 4D.**
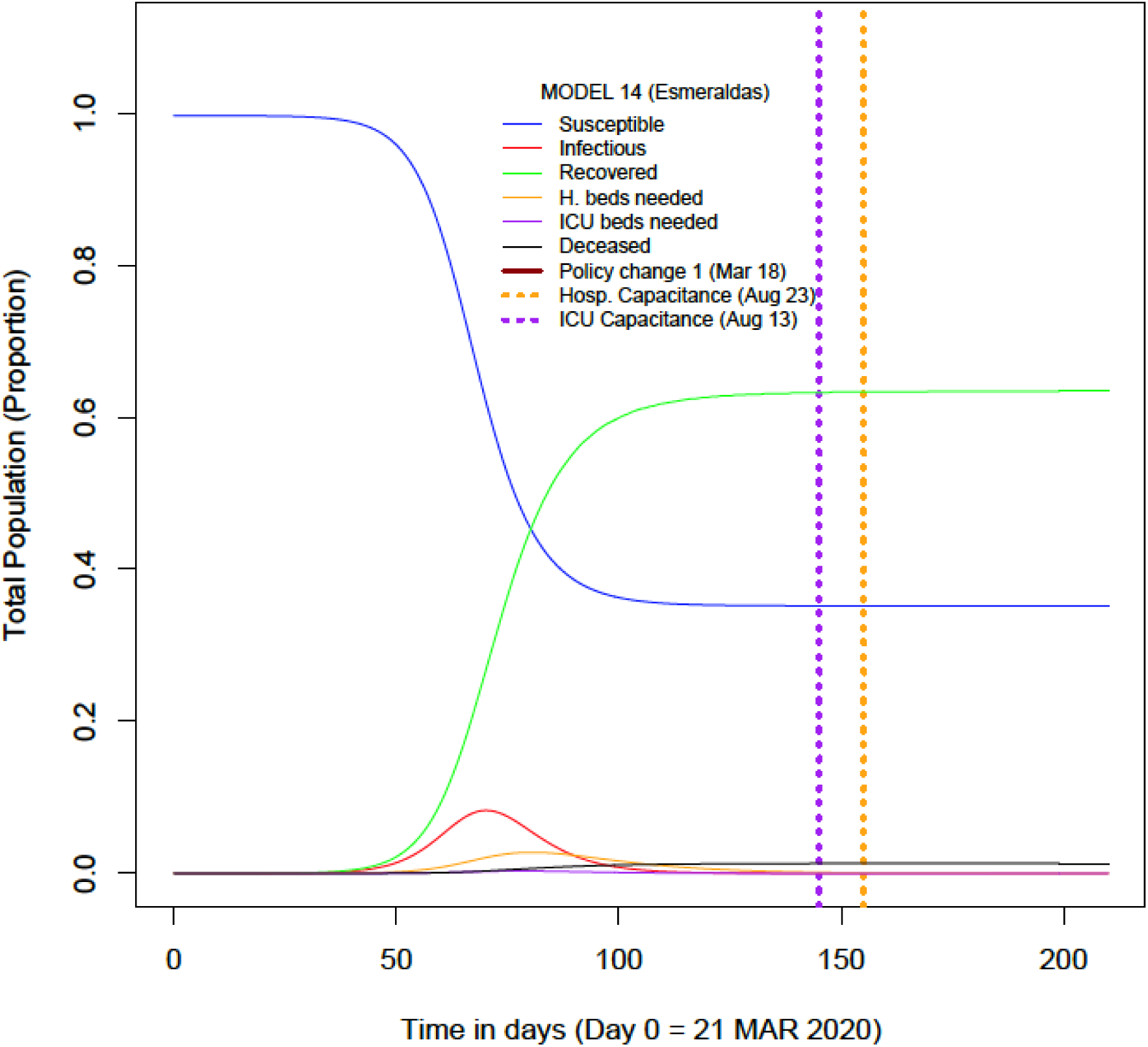
MODEL 14: HeCEM-simulated COVID-19 outbreak in Esmeraldas. Dashed lines represent time at which capacitance is reached.

We also explored the use of HeCEM in a city where the COVID-19 outbreak started after the enactment of containment measures. The first reported COVID-19 case was reported on March 21^st^ in Esmeraldas. (23) According to HeCEM (Figure 4D, Model 14), ICU-C would be reached by August 13 ^th^ whereas HC would be reached 10 days later, by August 23^rd^. By May 10^th^, the model predicts that 4,814 people have been infected since the beginning of the outbreak and that 50 people have died of COVID-19. The model also predicts that 32 healthcare workers would have been infected with COVID-19 by May 10^th^.

### Forecasting COVID-19 reported cases

A differenced first-order autoregressive ARIMA model was fit to forecast the cumulative COVID-19 incidence. The model was allowed to forecast up to July 25^th^. According to the forecasting, reported cases will continue an upward trend until reaching a total of 52,404 reported cases on July 25^th^ (mean = 21,343, SD = 16,9830; Figure 5).

**Figure 5.**
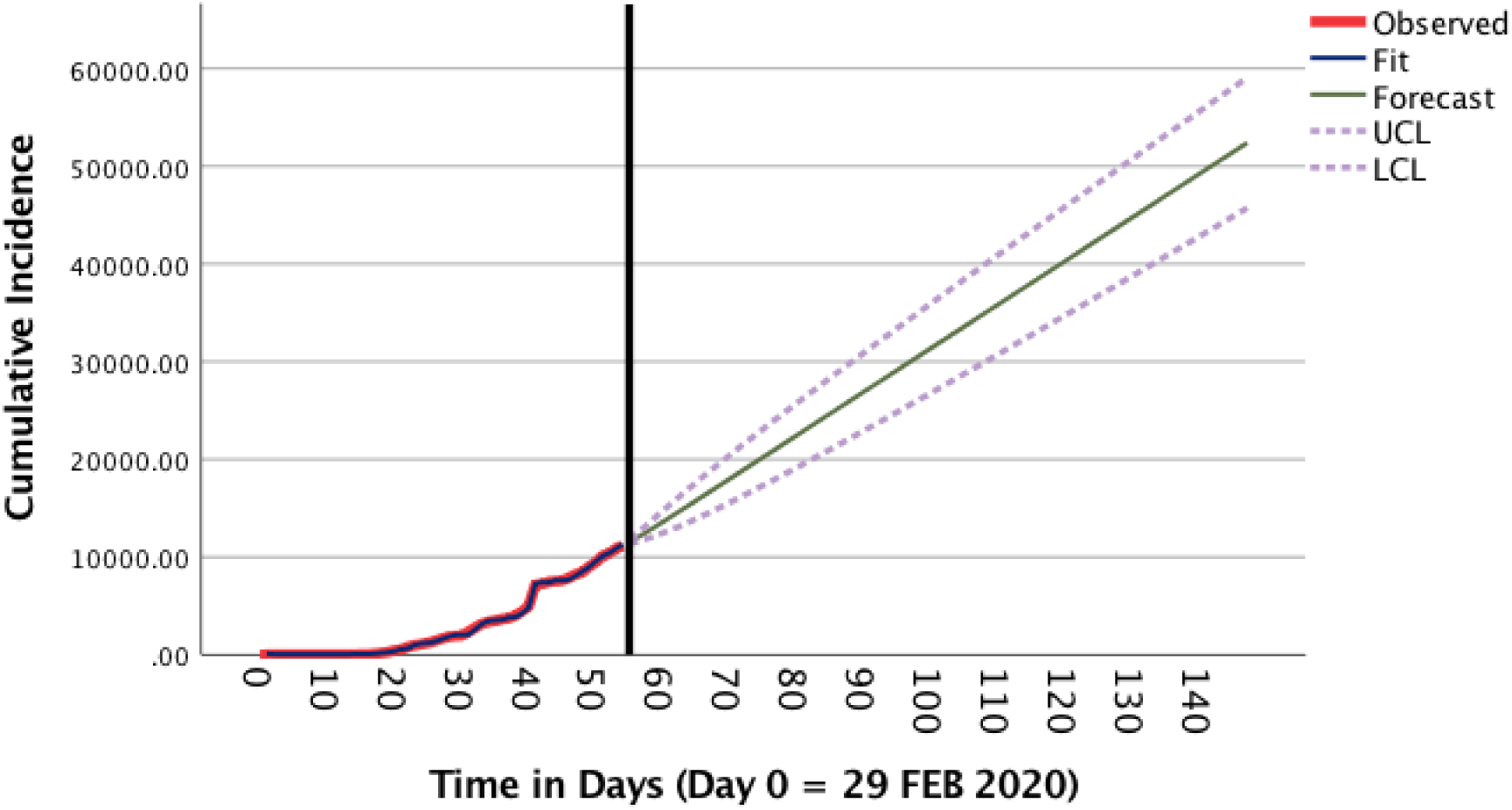
Forecast of COVID-19 cumulative reported incidence.

### Estimating simulation and predictive errors

ARIMA model-generated errors were the most stable, at around −0.05 (Figure S9). SIR model (Model 2) errors tend to decrease whereas HeCEM (Model 11) errors increase with time (Figure S9). SIR-generated errors were larger than HeCEM-generated errors (Table S6). ARIMA model-generated errors were not different than zero (Table S6). HeCEM-generated standardized errors were not different than the mean (p = 0.446, One-sample Wilcoxon Signed Rank test; Table S7).

## DISCUSSION

Based on reported figures, Ecuador has one of the largest COVID-19 outbreaks in the Americas, (1) and the largest in Latin America (in terms of ratios). However, the official figures are widely believed to underrepresent the true extent of the outbreak. (24) Guayaquil, in particular, has suffered the greatest burden in the country both in terms of morbidity and mortality. (25) The magnitude of the outbreak in Guayaquil has been dramatic. There have been reports of corpses accumulating on city streets after the local healthcare system was unable to cope with the demand and due to the overwhelming of morgues and funeral homes. (26)

Severe social distancing measures consisting on mobility restrictions were applied nationwide since March 18^th^, 2020. (5) Canton governments have been authorized to evaluate conditions to reverse containment measures. (6) Some cantons are considering lifting restrictions,(27) including Guayaquil that will keep current containment measures until May 24^th^. (28) We used SIR and HeCEM modeling to provide evidence that can inform decisions regarding reversing COVID-19 containment measures in Ecuador.

The SIR and HeCEM models reported in this manuscript derive transmission and infectiousness parameters from previous research studying COVID-19 transmission before and after the implementation of containment measures, including restrictions in mobilization, in China. (13) In addition, HeCEM models derive parameters from published literature as shown in Table 1. Since lockdown efforts in Ecuador seem to have been loosely implemented, at least in some regions,(29) it is likely that our estimates are rather conservative.

Our results suggest that Ecuador benefited from imposing containment measures by reducing cumulative COVID-19 mortality by over 130,000. Of all scenarios evaluated, the best outcome is given by delaying the implementation of a partial reversal of measures until both Hospital Capacitance and ICU Capacitance dates, under current circumstances, have been reached and stabilized. That corresponds to SIR MODEL 10 (i.e. partial reversal implemented on July 19^th^, 2020). HeCEM models may provide more accurate predictions, as they account for hospitalization and ICU admittance in addition to recovery and death. HeCEM models confirm that partial reversal of containment measures should occur after, or right before, HC and ICU-C have been reached. The HeCEM Guayaquil model predicts over 1.1 million cumulative infections by May 10^th^, 2020. A recent study conducted in Guayaquil suggests that over 30% of the population (close to 1 million people) have already been infected. (28) By April 30^th^, the HeCEM Guayaquil model estimated 7,299 deaths, which correlates with the excess mortality observed in Guayaquil (i.e. compared to 2019, there was an excess of 10,387 deaths between March and April, as registered by the Civil Registry). (30) SIR-generated errors were larger than HeCEM-generated errors suggesting that HeCEM models were more accurate. However, HeCEM-generated errors showed a tendency to increase over time. Estimating parameters from data generated on the field, in Ecuador, will help to reduce both SIR and HeCEM errors and produce more accurate predictions.

Reversing containment measures should be carefully considered. The WHO has indicated that COVID-19 cases may surge after lifting restrictions. (31) If Ecuador follows a similar trajectory, then the resulting surge in cases will be additional burden to an already overwhelmed healthcare system. Preventing overwhelming healthcare services will be key to handling post-pandemic SARS-CoV-2 transmission. (32) Prevention and control of cases in Ecuador, and specifically in densely-populated cities like Guayaquil, could be crucial to keeping the pandemic under control.

There are some limitations of this study. First, the study simulates the outbreak for the whole nation. It is important to keep in mind that in-country variations might exist. Similar approaches should be done at a local scale – we have started by providing two local HeCEM models. Second, the models assume that once a person recovers, they will not be re-infected. There is limited evidence regarding lifelong COVID-19 immunity. (33) Third, our models do not consider the socioeconomic impact of containment measures and therefore it does not include them in the analysis of the impact of reversing such measures. In terms of forecasting, the ARIMA models are based on the officially reported figures, which are believed to be an underrepresentation of the real number of cases. (24)

## Data Availability

Publicly available as cited.

## Acknowledgements

The authors would like to thank Dr. Catalina Lopez-Quintero for her valuable input.

## References

1. World Health Organization. Coronavirus disease (COVID-19). Situation Report 111. Available at: https://www.who.int/docs/default-source/coronaviruse/situation-reports/2020051Ocovid-19-sitrep-111.pdf?sfvrsn=1896976f_2. WHO [Geneva], May 10, 2020. Cited May 10, 2020.

2. Gobierno de la República del Ecuador. Secretaría General de Comunicación de la Presidencia. Se registra el primer caso de coronavirus en Ecuador [on line]. https://www.comunicacion.gob.ec/se-registra-el-primer-caso-de-coronavirus-en-ecuador/. Secretaría de Comunicación [Quito]. Cited April 29, 2020.

3. Gobierno de la República del Ecuador. Comité de Operaciones de Emergencia Nacional. Informe de Situación COVID-19 Ecuador No. 001 | COE Nacional −13 de Marzo de 2020. Available at: https://www.gestionderiesgos.gob.ec/wp-content/uploads/2020/03/Informe-de-Situaci%C3%B3n-No001-Casos-Coronavirus-Ecuador-12032020.pdf. COE Nacional [Quito], March 13, 2020. Cited, May 10,2020.

4. Gobierno de la República del Ecuador. Comité de Operaciones de Emergencia Nacional. Informe de Situación COVID-19 Ecuador No. 007 | COE Nacional −16 de Marzo de 2020 [on line]. Available at: https://www.gestionderiesgos.gob.ec/wp-content/uploads/2020/03/Informe-de-Situaci%C3%B3n-No007-Casos-Coronavirus-Ecuador-16032020-14h00.pdf. COE Nacional [Quito], March 16, 2020. Cited, May 10, 2020.

5. Gobierno de la República del Ecuador. Comité de Operaciones de Emergencia Nacional. Informe de Situación COVID-19 Ecuador No. 008 | COE Nacional -16 de Marzo de 2020 [on line]. Available at: https://www.gestionderiesgos.gob.ec/wp-content/uploads/2020/03/Informe-de-Situaci%C3%B3n-No008-Casos-Coronavirus-Ecuador-16032020-20h00.pdf. COE Nacional [Quito], March 16, 2020. Cited, May 10, 2020.

6. Gobierno de la República del Ecuador. Comité de Operaciones de Emergencia Nacional. Informe de Situación COVID-19 Ecuador No. 042 | COE Nacional - 29 de Abril de 2020. Available at: https://www.gestionderiesgos.gob.ec/wp-content/uploads/2020/04/Informe-de-Situaci%C3%B3n-No042-Casos-Coronavirus-Ecuador-29042020.pdf. COE Nacional [Quito], April 29, 2020. Cited, May 10,2020.

7. The World Bank. World Bank Open Data. Data [online]. https://data.worldbank.org/. The World Bank [Washington, D.C.] c. 2019. Cited April 29, 2020.

8. Institute Nacional de Estadística. Proyecciones Poblacionales. Institute Nacional de Estadística y Censos [online]. https://www.ecuadorencifras.gob.ec/proyecciones-poblacionales/. INEN [Quito]. Cited April 29, 2020.

9. Servicio Nacional de Gestión de Riesgos y Emergencias. COE Nacional [on line], https://www.gestionderiesgos.gob.ec/coe-nacional/. [Quito]. Cited April 29, 2020.

10. Soetaert K, Petzoldt T, Setzer RW. Solving Differential Equations in R: Package deSolve. 2010. 2010;33(9):25.

11. R Core Team (2020). R: A language and environment for statistical computing. R Foundation for Statistical Computing, Vienna, Austria. URL https://www.R-project.org/. Cited April 29, 2020.

12. Huang Y, Yang L, Dai H, Tian F & Chen K. Epidemic situation and forecasting of COVID-19 in and outside China. [Submitted]. Bull World Health Organ. E-pub: 16 March 2020. doi: http://dx.doi.org/10.2471/BLT.20.255158.

13. Li R, Pei S, Chen B, Song Y, Zhang T, Yang W, et al. Substantial undocumented infection facilitates the rapid dissemination of novel coronavirus (SARS-CoV2). Science. 2020.

14. Gobierno de la República del Ecuador. Instituto Nacional de Estadísticas y Censos. Camas y egersos hospitalarios [on line]. https://www.ecuadorencifras.gob.ec/camas-y-egresos-hospitalarios/. INEN [Quito], n.d. Cited, May 6, 2020.

15. Team CC-R. Severe Outcomes Among Patients with Coronavirus Disease 2019 (COVID-19) - United States, February 12-March 16, 2020. MMWR Morb Mortal Wkly Rep. 2020;69(12):343–6.

16. Verity R, Okell LC, Dorigatti I, Winskill P, Whittaker C, Imai N, et al. Estimates of the severity of coronavirus disease 2019: a model-based analysis. Lancet Infect Dis. 2020.

17. Garg S, Kim L, Whitaker M, O’Halloran A, Cummings C, Holstein R, et al. Hospitalization Rates and Characteristics of Patients Hospitalized with Laboratory-Confirmed Coronavirus Disease 2019 - COVID-NET, 14 States, March 1–30, 2020. MMWR Morb Mortal Wkly Rep. 2020;69(15):458–64.

18. Huang C, Wang Y, Li X, Ren L, Zhao J, Hu Y, et al. Clinical features of patients infected with 2019 novel coronavirus in Wuhan, China. Lancet. 2020;395(10223):497–506.

19. Wu Z, McGoogan JM. Characteristics of and Important Lessons From the Coronavirus Disease 2019 (COVID-19) Outbreak in China: Summary of a Report of 72314 Cases From the Chinese Center for Disease Control and Prevention. JAMA. 2020.

20. Gobierno de la República del Ecuador. Instituto Nacional de Estadísticas y Censos. Actividades y Recursos de Salud [on line]. Available at: https://www.ecuadorencifras.gob.ec/actividades-y-recursos-de-salud/. INEN [Quito], n.d. Cited, May 6, 2020.

21. IBM Corp. Released 2017. IBM SPSS Statistics for Windows, Version 25.0. Armonk, NY: IBM Corp.

22. Heredia, V, Gonzalez, J. Murió la primera paciente que dio positivo a covid-19 en Ecuador, informó el MSP; hay tres nuevos casos. https://www.elcomercio.com/actualidad/murio-primera-paciente-coronavirus-ecuador.html. March 13, 2020. Accessed April 30, 2020.

23. Gobierno de la República del Ecuador. Comité de Operaciones de Emergencia Nacional. Informe de Situación COVID-19 Ecuador No. 016 | COE Nacional - 21 de Marzo de 2020 [on line]. Available at: https://www.gestionderiesgos.gob.ee/wp-content/uploads/2020/03/Informe-de-Situaci%C3%B3n-No016-Casos-Coronavirus-Ecuador-21032020.pdf. COE Nacional [Quito], Mar 21, 2020. Cited, May 10,2020.

24. Otis J. COVID-19 Numbers Are Bad In Ecuador. The President Says The Real Story Is Even Worse. https://www.npr.org/sections/goatsandsoda/2020/04/20/838746457/covid-19-numbers-are-bad-in-ecuador-the-president-says-the-real-story-is-even-wo. National Public Radio, April 20, 2020. Cited May 14, 2020.

25. Gobierno de la República del Ecuador. Comité de Operaciones de Emergencia Nacional. Infografía No 073 C0VID 19 | COE Nacional -10 de Mayo de 2020 [on line]. Available at: https://www.gestionderiesgos.gob.ec/wp-content/uploads/2020/03/Resoluciones-COE-Nacional-24-de-marzo-2020.pdf. COE Nacional [Quito], May 10, 2020. Cited, May 10, 2020.

26. Rivers M, Gallón N. Where are the bodies? Missing remains mean no peace for grieving families in Ecuador [on line]. Cable News Network. May 10, 2020. https://www.cnn.com/2020/05/07/americas/ecuador-coronavirus-missing-intl/index.html. Cited May 10, 2020.

27. Medina A. 166 cantones se quedarán con semáforo en rojo hasta el 31 de mayo. El Comercio. https://www.elcomercio.com/actualidad/cantones-semaforo-rojo-covid19-distanciamiento.html. May 5, 2020. Accessed May 6, 2020.

28. El Universo. Coronavirus en Guayaquil: Semáforo en rojo se mantendra hasta el 24 de mayo; según muestreo, el 33% de la población se ha contagiado, de esos el 14,8% ha superado la enfermedad [on line]. Available at: https://www.eluniverso.com/guayaquil/2020/05/07/nota/7834590/guayaquil-coronavirus-medidas-coe. El Universo [Guayaquil], May 7, 2020. Cited May 10, 2020.

29. Torres I, Sacoto F. Localising an asset-based COVID-19 response in Ecuador. Lancet. 2020;395(10233):1339.

30. Ecuavisa. Muertos sin descanso | Visión 360 VII Temporada [video, on line]. Available: https://www.youtube.com/watch?v=gXP2eJW5hnA. Ecuavisa [Quito], May 11, 2020. Cited May 12, 2020.

31. World Health Organization. WHO Director-General’s opening remarks at the media briefing on COVID-19-11 May 2020 [on line]. https://www.who.int/dg/speeches/detail/who-director-general-s-opening-remarks-at-the-media-briefing-on-covid-19---11-may-2020. WHO [Geneva], May 11, 2020. Cited May 12, 2020.

32. Kissler SM, Tedijanto C, Goldstein E, Grad YH, Lipsitch M. Projecting the transmission dynamics of SARS-CoV-2 through the postpandemic period. Science. 2020.

33. Kirkcaldy RD, King BA, Brooks JT. COVID-19 and Postinfection Immunity: Limited Evidence, Many Remaining Questions. JAMA. 2020.

